# Diagnostic accuracy of Clinical Radiology Reports for Trauma Radiographs: A retrospective validation study

**DOI:** 10.1101/2025.07.16.25331604

**Authors:** Frederik Jager Bruun, Felix C. Müller, Janus Uhd Nybing, Dimitar Ivanov Radev, Amir Roshani Honar, Jimi Ravn Preuthun, Frederikke Walther Laurberg, Mathias Willadsen Brejnebøl

## Abstract

**Background/purpose:** Development and validation of AI tools for diagnostic imaging requires high-quality annotations. Dedicated research readings are considered superior to the clinical radiologic report (CRR). However, the practices for generating CRR varies in rigor. The purpose of this study was to validate CRRs produced using a post-conference multiple disciplinary reader workflow by comparing them to dedicated research readings of trauma radiographs.

**Materials and Methods:** This retrospective study included consecutive patients referred for radiography at the indication of trauma at two university hospitals. For the index test, the CRRs were evaluated based on their description of eight common diagnoses. The reference standard was established as the agreement of two certified reporting technologists and arbitrated by a senior musculoskeletal radiologist. Sensitivity, specificity, positive and negative predictive values were calculated.

**Results:** The study sample consisted of 618 consecutive examinations (median age 52 years IQR, 24, 76, 351 female). Fracture incidence was 36%. Incidence of other findings ranged from 1% (bone lesions) to 10% (degenerative disease).

The sensitivities of the CRRs were: fracture (97% [94 to 99%]), luxation (87% [69 to 96%]), degenerative disease (67% [53 to 78%]), effusion (67% [46 to 83%]), old fracture (64% [48 to 78%]), subluxation (44% [14 to 79%]), halisteresis (30% [13 to 53%]) and bone lesion (25% [1 to 81%]). Specificity ranged from halisteresis (94% [92 to 96%]) to subluxation (100% [99 to 100%]) and luxation (100% [99 to 100%]).

**Conclusion:** We found that the workflow at the tested hospitals generates clinical radiologic reports of trauma radiographs with a diagnostic accuracy for the assessment of fracture and luxation suitable for research quality labelling.

## Introduction

Development and validation of Artificial intelligence (AI) tools for diagnostic imaging require high-quality annotation of relevant clinical examinations. Dedicated research readings are recognised as the most rigorous way of obtaining such annotations. Because dedicated readings are costly, the clinical radiology reports (CRR) are often used instead. However, the practices for generating CRRs vary in rigor across examination types and departments. In light of these considerations, the purpose of this study is to validate findings extracted from CRR against dedicated research readings of trauma radiographs.

Well annotated imaging data for development of AI tools is scarce(1). It is suggested that monitoring of clinically employed AI tools should include, at minimum, yearly re-evaluations(2), that rely on high-quality ground truth information(3). Using readings already done by experts is a convenient way of obtaining this information that does not require radiology experts to divert time from clinical practice. However, if the CRRs lack sufficient completeness or diagnostic precision, their usage carries the risk of introducing systematic misclassification otherwise known as information bias(4).

Perceptual errors such as underreading and satisfaction of search remain the most frequent causes of diagnostic mistakes in radiology(5) (6). Additionally, to keep a CRR relevant clinically insignificant findings are often omitted(7). In research settings, these errors and omissions are mitigated through multiple readings and a strict labelling scheme. In clinical practice, institutions may rely on double readings, pre-reading by trainees, or discussion at clinical conferences. The way institutions allocate these measures differs between examinations, resulting in varying quality of CRRs.

Although data validation studies are increasingly common in epidemiologic literature(8), sometimes with dedicated publication categories(9), such methodological scrutiny remains rare in AI-focused imaging research.

It is well known that different diagnoses are associated with higher interreader variability than others(10) (11) (12). For some diagnoses, the interreader variability is related to the suitability of the imaging modality used. However, we hypothesise that diagnostic accuracy of the CRR is also influenced by the clinical relevance of the suspected diagnosis to the indication for imaging referral. Our primary aim was to identify common findings in clinical radiologic reports of trauma radiographs with a diagnostic accuracy suitable for research quality labelling.

## Materials and Methods

The need for informed written consent was waived by the institutional review board (ID 22070206) due to the retrospective nature of the study. The manuscript was prepared according to the STARD (STandards for Reporting of Diagnostic Accuracy Studies) guidelines(13).

### Study sample

The study sample consisted of all consecutive radiographs of patients referred from the emergency departments of two university hospitals on the basis of trauma. The radiographs from Herlev hospital were acquired from January 08 to 17, 2023, and the radiographs from Bispebjerg hospital were acquired from January 01 to 09, 2023. Patients were identified in November 2023 by searching the local PACS (Impax 6; Agfa HealthCare). No examinations were excluded.

### Index test

At our institutions bone radiographs are read by reporting technologists. Trauma radiographs at our institutions are assessed by the emergency physician, at least one reporting technologist, and usually also an in-training radiologist before it is discussed at a conference with a senior orthopaedic surgeon. Supplementary imaging is typically performed prior to the conference. Thereafter the CRR is finalised. If an error is discovered later, it is possible to write an addition correcting the finalised CRR. Due to the retrospective nature of the study, the CRR was finalised before the reference standard was established. See Figure 1 for illustration of this process.

**Figure 1:**
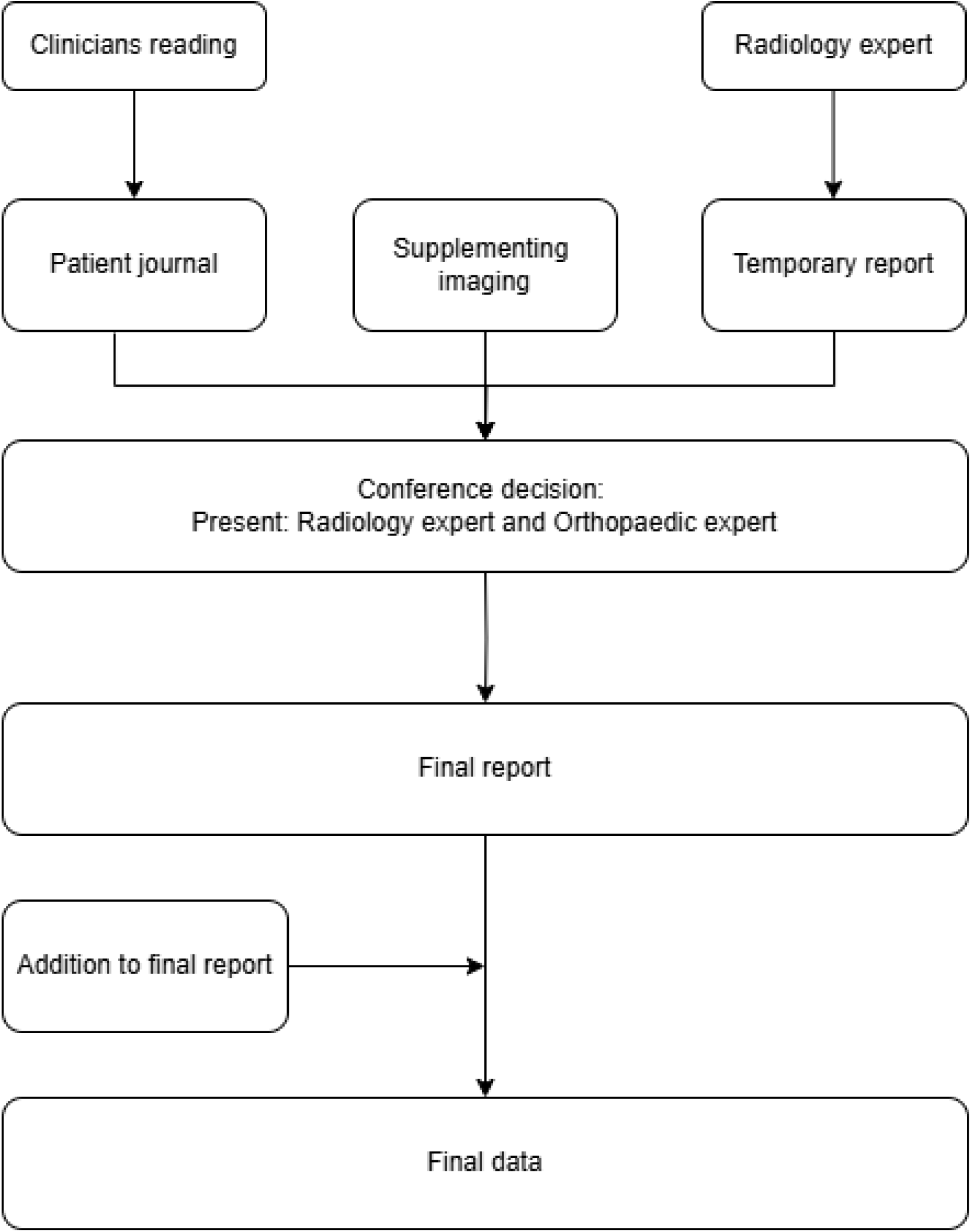
Diagram illustrating the process of generating a clinical radiologic report for trauma radio-graphs at the hospitals participating in the study.

**Figure 2:**
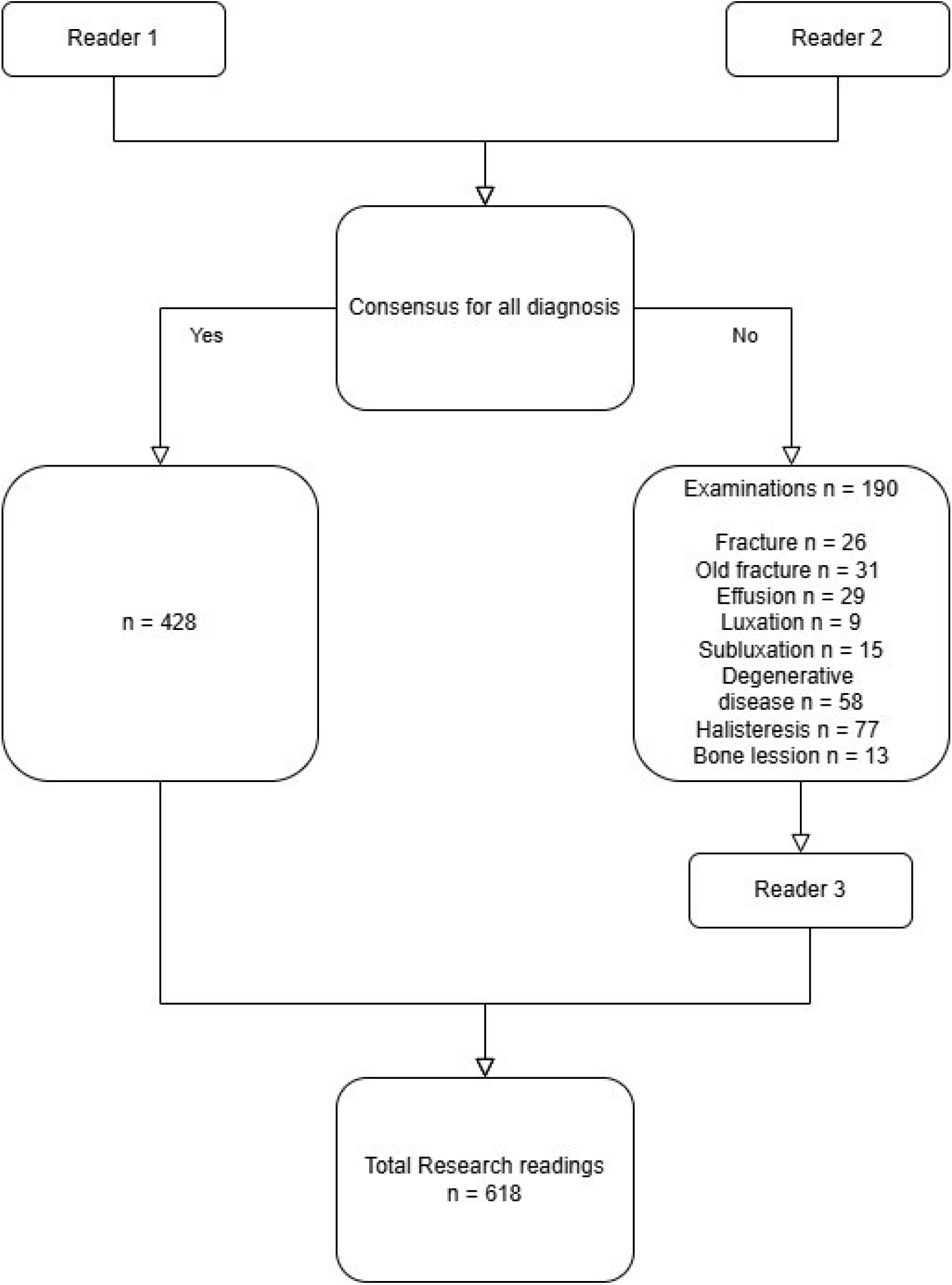
Flowchart indicating the process of generating the research readings.

The finalised CRRs were annotated by a physician with 3 years radiology experience (F.J.B.) and labelled in REDCAP(14). The reports were assessed for description of these typical findings on bone radiographs: fracture, old fracture, sub luxation, luxation, effusion, bone lesion, degenerative disease and halisteresis. Findings were annotated dichotomously as either present or absent. In cases where a finding was mentioned as uncertain, it was annotated as present. If a finding was not explicitly mentioned, it was considered absent from the report.

### Reference standard

The examinations were double read by a team of four reporting technologists (A.R.H, J.R.P, F.W.L with 4-11 years of experience as reporting technologists). All four have undergone two years post graduate education in reading trauma radiographs and perform this task as their primary clinical responsibility. A meta-analysis has shown that trained reporting technologists achieved the same diagnostic accuracy as radiologist for reporting trauma radiographs(15).

For establishing the reference standard, there was access to prior clinical reports, clinician’s referral notes, patients’ clinical journal, CRR and both historic and follow up imaging. There was at least 15 months of follow-up time at the time of establishing the reference standard.

To eliminate time pressure, each technologist was given two full workdays to complete readings at their own pace. No minimum number of cases was required. A computer program (Redcap) ensured that examinations were only read twice. Reference readers were instructed to annotate the presence of the eight findings if they considered it appropriate to report in a trauma reporting setting. Discrepancies were settled by D.I.R., a musculoskeletal radiologist with 20 years of experience.

### Statistical Analysis

Sensitivity, specificity, positive predictive value, and negative predictive value with 95% confidence intervals (CI) were calculated using the binomial exact method. A two-sample test for equality of proportions was used to test diagnostic accuracy across hospitals. Cohen’s κ values and disagreement fractions were calculated to assess interreader variability among the reference readers. Reader identity was not part of the scope of the analysis so to minimise risk of reader bias, the order of the two readings per case was randomised.

Statistical analyses were carried out by F.J.B. using R Software (v.4.3.0). Code for metrics calculation can be seen in the supplementary. The packages binom (v.1.1.1.1) and psych (v.2.4.6.26) were used for confidence interval calculation.

## Results

The study sample consisted of 618 consecutive examinations from Bispebjerg and Herlev university hospitals. The median age of the patients was 52 years (IQR 24-76). There were 351 (57%) female patients. The incidence of fractures was 36% resulting in 225 reports describing this finding. The anatomical location of the fractures was ankle 24, clavicula 4, elbow 4, foot 27, hand 36, hip 40, knee 10, shoulder 22, wrist 42 and other 16. The incidence of other diagnoses ranged from 1% (4/618) for bone lesions to 10% (60/618) for degenerative disease.

Measures of diagnostic accuracy for the individual diagnoses are presented in Table 2 and Figure 3 and cases of disagreement between research readings and the CRR are presented in Figure 4. We found the highest sensitivity for fractures (97% [94 to 99%] 219/225) and second highest for luxation (87% [69 to 96%] 26/30). Degenerative disease (67% [53 to 78%] 40/60), effusion (67% [46 to 83%] 18/27) and old fracture (64% [48 to 78%] 27/42) had almost the same sensitivity. The lowest sensitivities were seen for subluxation (44% [14 to 79%] 4/9), halisteresis (30% [13 to 53%] 7/23) and bone lesion (25% [1 to 81%] 1/4).

**Figure 3:**
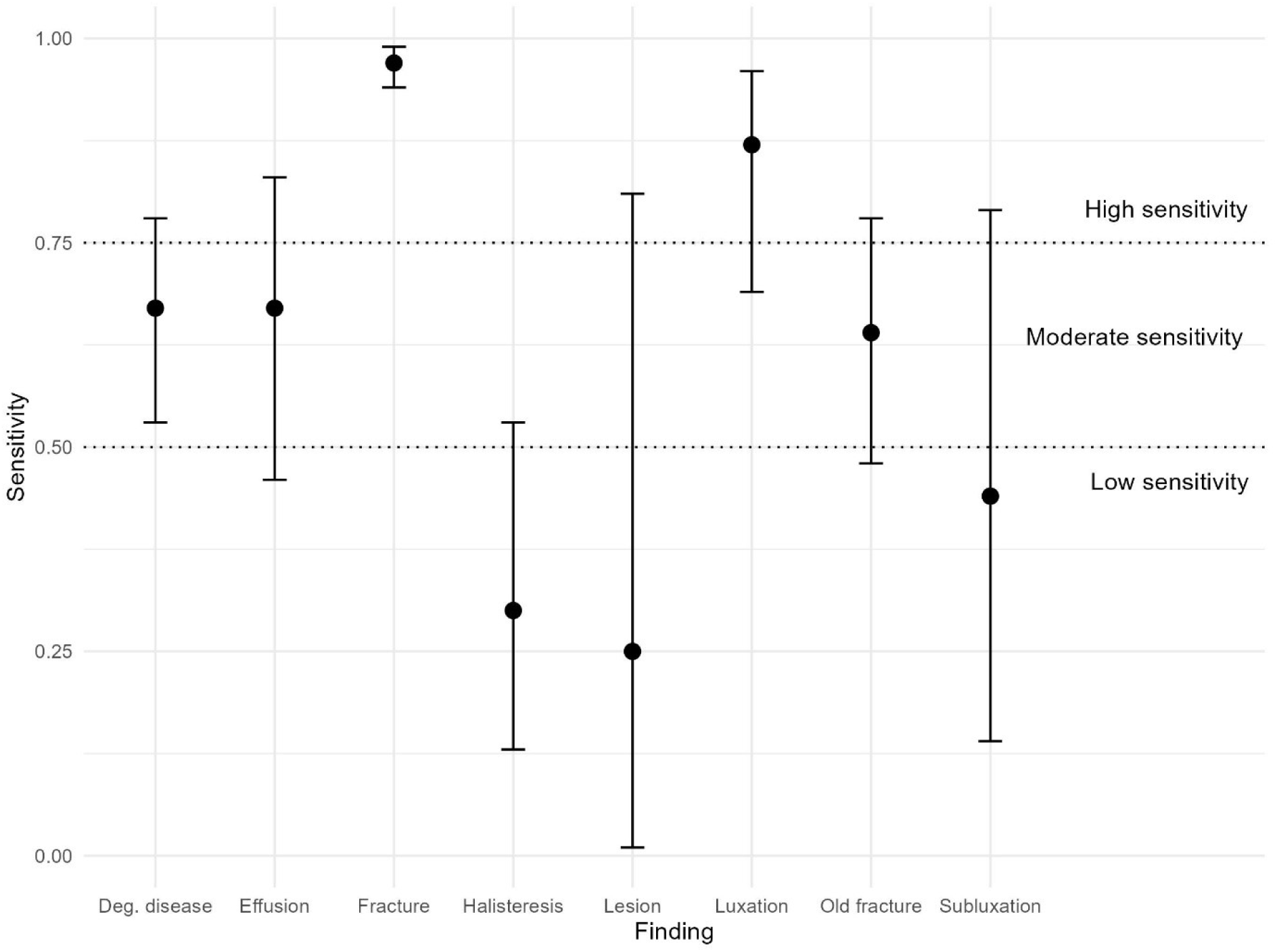
Illustrates the sensitivity with 95% confidence intervals of each of the eight findings of the trauma radiographs.

**Figure 4:**
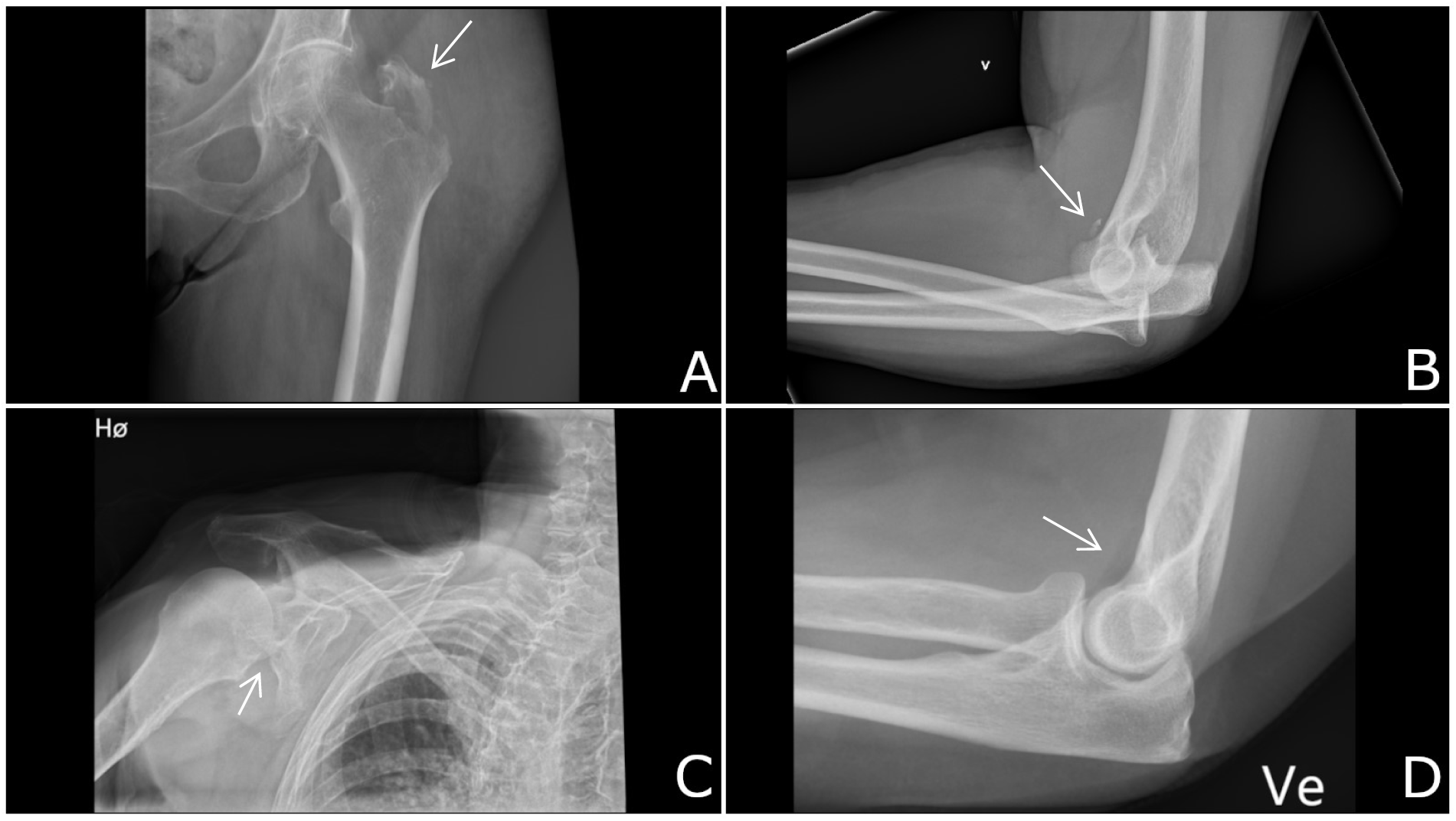
Cases of disagreement between research readings and the CRR. A: Female in her late eighties presenting with pain, hematoma in the left hip area, and weak pulses and cyanosis in both feet after a fall. The CRR describes an avulsion fracture of the greater trochanter, with 1 mm cranial and medial displacement. The research readings did not confirm the presence of a fresh fracture. B: Male in his late fifties arrives for his second radiograph with an elbow dislocation that is not fully reduced. The CRR doesn’t mention an avulsion of the coronoid process. However, this finding was mentioned at his first examination earlier that day. C: Female in her early nineties presenting with pain and restricted movement of the right upper extremity following a high-speed fall from a scooter. The CRR describes no fractures or dislocations. Three days later, a CT scan revealed a scapular fracture, which was also identified in the research readings. D: Female in her late seventies presenting with pain localized to the lateral humeral epicondyle and radial head following a fall. The CRR describes a small effusion of the elbow joint. However, the research readings disagreed with this finding.

**Table 1:** Patient characteristics.

|  | Overall<br>n = 618<sup>1</sup> | Herlev<br>hospital<br>N = 412<sup>1</sup> | Bispebjerg<br>hospital<br>n = 206<sup>1</sup> |
| --- | --- | --- | --- |
| Sex |  |  |  |
| Female | 351 (57%) | 232 (56%) | 119 (58%) |
| Male | 267 (43%) | 180 (44%) | 87 (42%) |
| Child | 106 (17%) | 103 (25%) | 3 (1.5%) |
| Age | 52 (24, 76) | 49 (18, 74) | 59 (35, 79) |
| Fracture | 225 (36%) | 145 (35%) | 80 (39%) |
| Old fracture | 42 (6.8%) | 20 (4.9%) | 22 (11%) |
| Subluxation | 9 (1.5%) | 8 (1.9%) | 1 (0.5%) |
| Luxation | 30 (4.9%) | 18 (4.4%) | 12 (5.8%) |
| Effusion | 27 (4.4%) | 18 (4.4%) | 9 (4.4%) |
| Halisteresis | 23 (3.7%) | 9 (2.2%) | 14 (6.8%) |
| Degenerative disease | 60 (9.7%) | 27 (6.6%) | 33 (16%) |
| Lesion | 4 (0.6%) | 3 (0.7%) | 1 (0.5%) |
| <sup>1</sup> n (%); Median (Q1, Q3) |  |  |  |

**Table 2:** Diagnostic Accuracy Metrics for individual diagnosis.

| Finding | TP | TN | FN | FP | Sensitivity (95% CI) | Specificity (95% CI) | PPV (95% CI) | NPV (95% CI) |
| --- | --- | --- | --- | --- | --- | --- | --- | --- |
| Fracture | 219 | 382 | 6 | 11 | 0.97 (0.94–0.99) | 0.97 (0.95–0.99) | 0.95 (0.92–0.98) | 0.98 (0.97–0.99) |
| Old fracture | 27 | 564 | 15 | 12 | 0.64 (0.48–0.78) | 0.98 (0.96–0.99) | 0.69 (0.52–0.83) | 0.97 (0.96–0.99) |
| Subluxation | 4 | 606 | 5 | 3 | 0.44 (0.14–0.79) | 1 (0.99–1) | 0.57 (0.18–0.9) | 0.99 (0.98–1) |
| Luxation | 26 | 588 | 4 | 0 | 0.87 (0.69–0.96) | 1 (0.99–1) | 1 (0.87–1) | 0.99 (0.98–1) |
| Effusion | 18 | 578 | 9 | 13 | 0.67 (0.46–0.83) | 0.98 (0.96–0.99) | 0.58 (0.39–0.75) | 0.98 (0.97–0.99) |
| Halisteresis | 7 | 559 | 16 | 36 | 0.3 (0.13–0.53) | 0.94 (0.92–0.96) | 0.16 (0.07–0.31) | 0.97 (0.96–0.98) |
| Degenerative disease | 40 | 531 | 20 | 27 | 0.67 (0.53–0.78) | 0.95 (0.93–0.97) | 0.6 (0.47–0.72) | 0.96 (0.94–0.98) |
| Lesion | 1 | 605 | 3 | 9 | 0.25 (0.01–0.81) | 0.99 (0.97–0.99) | 0.1 (0–0.45) | 1 (0.99–1) |

We found the lowest specificity for halisteresis (94% [92 to 96%] 559/595) and the highest for subluxation (100% [99 to 100%] 606/606) and luxation (100% [99 to 100%] 588/588).

For the positive predictive value (PPV), luxation had a perfect score (100% [87 to 100%] 26/26). Fracture had the second highest PPV (95% [92 to 98%] 219/230). The group of diagnoses with intermediate PPV consisted of old fracture (69% [52 to 83%] 27/39), degenerative disease (60% [47 to 72%] 40/67), effusion (58% [39 to 75%] 18/31) and subluxation (57% [18 to 90%] 4/7). There were two diagnoses that had markedly lower PPV than the rest. These where halisteresis (16% [7 to 31%] 7/43) and bone lesion (10% [0 to 45%] 1/10)

We found the negative predictive value (NPV) to range from 96% for degenerative disease (95% CI: 94% to 98% 531/551) to 100% for bone lesion (95% CI: 99% to 100% 605/608).

Interhospital differences in diagnostic accuracy are presented in Table 3. For fracture detection, there was no evidence of a difference between hospitals in neither sensitivity (p = 0.91) nor specificity (p = 0.76). The only difference across hospitals in sensitivity was seen for luxation (p = 0.01; sensitivity: Herlev 100% [81 to 100%]; Bispebjerg, 67% [35 to 90%]). There was difference in specificity for five diagnoses: bone lesion (p = 0.03; Herlev 98% [96 to 99%]; Bispebjerg, 100% [98 to 100%]), old fracture (p = 0.01; Herlev 99% [97 to 100%]; Bispebjerg, 96% [92 to 98%]), subluxation (p = 0.01; Herlev 100% [99 to 100%]; Bispebjerg, 99% [96 to 100%]), halisteresis (p = < 0.001; Herlev 99% [97 to 99%]; Bispebjerg, 84% [78 to 89%]) and degenerative disease (p = < 0.001; Herlev 99% [97 to 100%]; Bispebjerg, 87% [81 to 92%]).

**Table 3:** Sensitivity and specificity stratified by hospital - z-test for difference in mean across hospitals.

|  | Sensitivity | Sensitivity | Z-test | Specificity | Specificity | Z-test |
| --- | --- | --- | --- | --- | --- | --- |
|  | Herlev hospital | Bispebjerg hospital |  | Herlev hospital | Bispebjerg hospital |  |
| **Fracture** | 0.97 (0.93–0.99)<br>[141 / 145] | 0.98 (0.91–1)<br>[78 / 80] | 0.91 | 0.97 (0.95–0.99)<br>[260 / 267] | 0.97 (0.92–0.99)<br>[122 / 126] | 0.76 |
| **Old fracture** | 0.5 (0.27–0.73)<br>[10 / 20] | 0.77 (0.55–0.92)<br>[17 / 22] | 0.07 | 0.99 (0.97–1)<br>[388 / 392] | 0.96 (0.92–0.98)<br>[176 / 184] | 0.01 |
| **Subluxation** | 0.5 (0.16–0.84)<br>[4 / 8] | 0 (0–0.98)<br>[0 / 1] | 0.34 | 1 (0.99–1)<br>[404 / 404] | 0.99 (0.96–1)<br>[202 / 205] | 0.01 |
| **Luxation** | 1 (0.81–1)<br>[18 / 18] | 0.67 (0.35–0.9)<br>[8 / 12] | 0.01 | 1 (0.99–1)<br>[394 / 394] | 1 (0.98–1)<br>[194 / 194] | 1 |
| **Effusion** | 0.72 (0.47–0.9)<br>[13 / 18] | 0.56 (0.21–0.86)<br>[5 / 9] | 0.39 | 0.98 (0.97–0.99)<br>[388 / 394] | 0.96 (0.93–0.99)<br>[190 / 197] | 0.11 |
| **Haliteresis** | 0.22 (0.03–0.6)<br>[2 / 9] | 0.36 (0.13–0.65)<br>[5 / 14] | 0.49 | 0.99 (0.97–0.99)<br>[397 / 403] | 0.84 (0.78–0.89)<br>[162 / 192] | <0.001 |
| **Degenerative disease** | 0.59 (0.39–0.78)<br>[16 / 27] | 0.73 (0.54–0.87)<br>[24 / 33] | 0.27 | 0.99 (0.97–1)<br>[380 / 385] | 0.87 (0.81–0.92)<br>[151 / 173] | <0.001 |
| **Lesion** | 0.33 (0.01–0.91)<br>[1 / 3] | 0 (0–0.98)<br>[0 / 1] | 0.5 | 0.98 (0.96–0.99)<br>[400 / 409] | 1 (0.98–1)<br>[205 / 205] | 0.03 |

Independent readings of the reference readers, before arbitration had a disagreement for at least one diagnosis between the two research readings in 31% of the examinations (190/618). Disagreement for individual diagnosis can be seen in Figure 2 and selected cases of disagreement between the reference readers are presented in Figure 5. The interreader agreement for the individual diagnosis is presented in Table 4. The highest Cohens κ score was seen for fracture (0.92 95% CI 0.89 to 0.95) and luxation (0.83 95% CI 0.71 to 0.94). We found moderate kappa scores for effusion (0.6 95% CI 0.46 to 0.73), degenerative disease (0.58 95% CI 0.48 to 0.68) and old fracture (0.53 95% CI 0.39 to 0.68). Lesion (−0,01 95% CI −0.01 to 0), halisteresis (0.33 95% CI 0.22 to 0.45) and subluxation (0.35 95% CI 0.09 to 0.61) had low to very low κ measures. The highest disagreement fractions were seen for Halisteresis (12%) and degenerative disease (9%), while the lowest were seen for luxation (1%), bone lesion (2%) and subluxation (2%). The intermediate group of disagreements consisted of fracture (4%), effusion (5%) and old fracture (5%)

**Figure 5:**
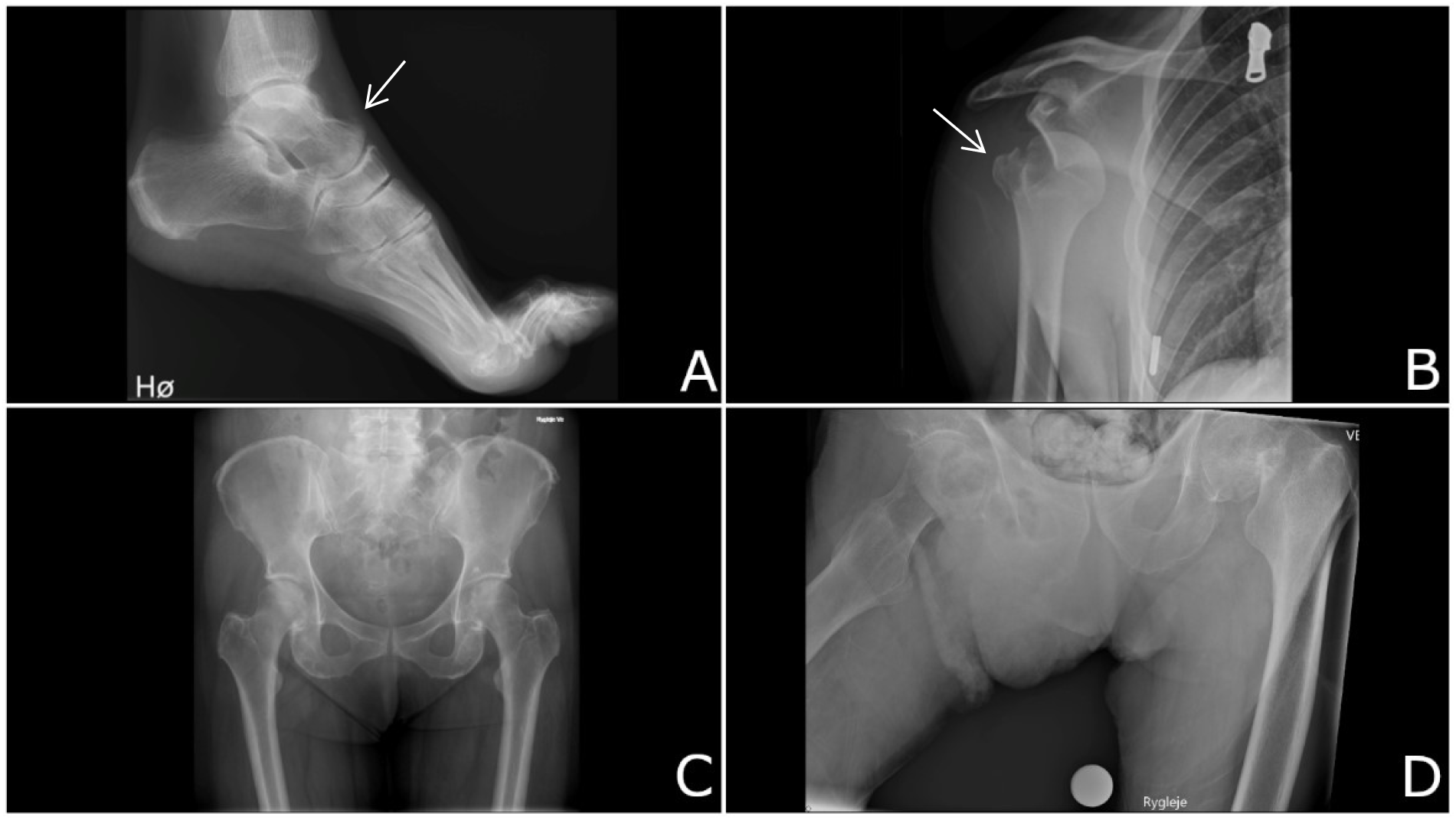
Cases of disagreement between the two reference readers of the research readings. A: Female in her late eighties with direct pain from the lateral malleolus. The radiographs show a small, slightly rounded fragment dorsal to the neck of the talus. The readers differed in whether this represented a fresh fracture or an old avulsion. B: Male in his early twenties presenting with a misaligned shoulder and severe pain. The right upper extremity is hanging and cannot be moved actively or passively without pain. The radiographs clearly show dislocation of the shoulder. It is likely that the disagreement between the expert readers relies on a data entry error. C: Female in her late sixties referred with the note: “obs. frac.” The expert readers of this radiograph differed on whether she had signs of degenerative disease. D: Male in his late eighties with suspected hip fracture and impaired cooperability had a radiograph of poor quality. The expert readers differed in opinion on the presence of halisteresis.

**Table 4:**
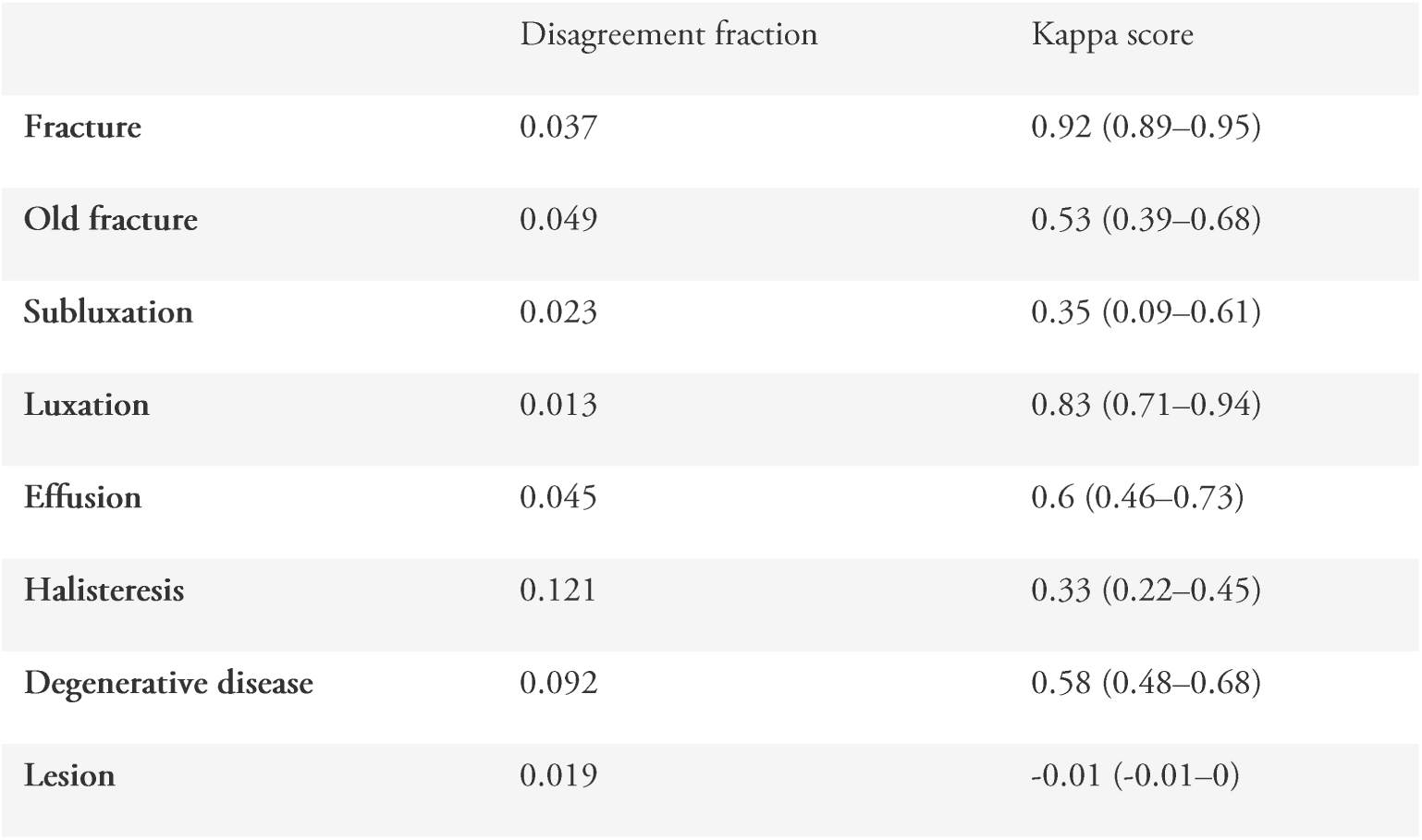
Interreader reliability of the reference standard by absolute disagrement fragmentions and Cohens κ scoring.

|  | Disagreement fraction | Kappa score |
| --- | --- | --- |
| Fracture | 0.037 | 0.92 (0.89–0.95) |
| Old fracture | 0.049 | 0.53 (0.39–0.68) |
| Subluxation | 0.023 | 0.35 (0.09–0.61) |
| Luxation | 0.013 | 0.83 (0.71–0.94) |
| Effusion | 0.045 | 0.6 (0.46–0.73) |
| Halisteresis | 0.121 | 0.33 (0.22–0.45) |
| Degenerative disease | 0.092 | 0.58 (0.48–0.68) |
| Lesion | 0.019 | -0.01 (-0.01–0) |

## Discussion

The clinical radiologic report is a readily available label for diagnoses, but concerns exist regarding its validity. In this retrospective validation study of the CRR of trauma radiographs we found that the sensitivity for the individual diagnosis were heterogenous ranging from 25% (bone lesion, 95%CI 1% to 81%) to 97% (fracture, 95%CI 94% to 99%). The specificities were all very high with the lowest value for halisteresis (94% 95%CI 92% to 96%).

The diagnostic accuracy of the CRR could be grouped into three categories, one group with very high diagnostic accuracy. One group with intermediate diagnostic accuracy and a group with a low diagnostic accuracy. For validating the clinical performance of AI tools against the CRR we can only recommend the diagnosis in the very high diagnostic accuracy group. In the category of almost perfect agreement between CRR and radiologic report we found fracture (sensitivity, 97% [93% to 99%]) and luxation (sensitivity, 87% [69 to 96%]). In the group of diagnosis with a moderate diagnostic accuracy the diagnosis is made consistently and the CRR can be used for detecting prevalence differences across populations but isn’t reliable for individual diagnosis. The group of diagnosis with moderate reliability of the CRR consisted off degenerative disease (sensitivity, 67% [53% to 78%]), effusion (sensitivity, 67% [46% to 83%]) and old fracture (sensitivity, 64% [48% to 78%]). When it comes to the group of diagnosis with the lowest diagnostic accuracy, we found the reliability to be so low that we advise against using the CRR for any validation studies based on these diagnoses. In the very low category, we found subluxation (sensitivity, 44% [14% to 79%]), halisteresis (sensitivity, 30% [13% to 53%]) and bone lesion (sensitivity, 25% [1% to 81%]).

Assessing interreader agreement showed a one-to-one correspondence between diagnostic accuracy and the κ score. This suggests that the validity of the CRR is low for the same findings as those where the reference readers struggle to agree. This supports that the amount of diagnostically uncertain patients for a given diagnosis is a major determinant of the CRR validity.

However, Cohen’s κ can produce paradoxically low values in settings with very low prevalence(16). To obtain a complete understanding of interreader agreement in low prevalence populations, κ must be interpreted in conjunction with a measure of marginal prevalence(17) (18) (19). Bone lesions had the second-lowest disagreement fraction, suggesting that this finding’s extremely low κ score is to some degree misleading. Halisteresis had the highest disagreement fraction and the second-lowest κ value, confirming that halisteresis is a highly uncertain finding. In contrast to the low prevalence findings, fracture had a prevalence of 36% and consequently yielded a very high κ score of 0.92, despite a disagreement fraction of 4%.

To keep a CRR relevant clinically insignificant findings are often omitted(7), this contrasts with research readings where one is required to take a stance on every finding. This leads research readings to produce more positive findings. Hence, negatively affecting the sensitivity of the CRRs in our study. Fracture and luxation are typically the main concern of the referring physician when referring for trauma radiography, these findings had a markedly higher diagnostic accuracy then the rest. The group of diagnosis for which the CRRs had very low diagnostic accuracy consisted of subluxation, halisteresis and bone lesion which are diagnosis that are never the main concern of a referral for trauma radiography. A study using CRRs from a sample of patients referred for assessment of degenerative disease would probably find a higher diagnostic accuracy for this diagnosis (20) (21). The threshold for when to call out a potentially serious finding varies by reader but is also adapted to the individual situation of a patient. Earlier experiences from our research groups studies of lung radiographs(22) and brain MRI(23) supports the view that CRR’s are of higher quality regarding diagnosis typically relevant for the clinical situation the patient is referred from.

We consider the generalisability of the findings in this study limited to other institutions that has a reporting setup as rigorous as our institutions and caution should be paid if the results are used where supplementing CT-scans isn’t as readily available as at the hospitals of this study. Secondly, the prevalence of several findings was low resulting in limited statistical power. A third limitation is that the expert readers had access to the CRR during their research readings. This could lead to an overestimation of the diagnostic accuracy of the CRRs. Fourth the pairing of readers for each examination was not controlled and resulted in a screwed distribution of pairings. This was a consequence of the liberation of time pressure by not requiring a specific number of readings from each reader.

In **conclusion** we found that the workflow at the tested hospitals generates clinical radiologic reports of trauma radiographs with a diagnostic accuracy for the assessment of fracture and luxation suitable for research quality labelling. With the current development of large language models’ ability to turn free text into structured data, high quality CRRs allow for automatic validation of artificial intelligence tools.

## Abbrevations

AI: artificial intelligence
CRR: Clinical radiologic report

## Data Availability

All data produced in the present study are available upon reasonable request to the authors.

## Notes

### Competing Interest Statement

The authors have declared no competing interest.

### Funding Statement

Salary for the main author was provided by the innovation fund denmark project: CLIC Clinical Imaging Consortium

### Author Declarations

The Hospital Executive Board of Bispebjerg and Frederiksberg Hospital, the Hospital Executive Board of Herlev and Gentofte Hospital and the Hospital Executive Board of north zealands hospitals gave waived ethical approval for this retrospective work. The work was categorized under quality control and development which doesn't require ethical aproval for access to patient data (Sundhedslovens paragraph 42). Aproval was given under WorkZone nr. 22070206 and stored by the administration of the capital region of denmark. Data was collected in a secure database (REDcap). After collection of data, the dataset was deidentified and exported for analysis in this study. This was done in conformity to the regulations in the aproval of the work.

